# Effectiveness of the third dose of BNT162b2 vaccine on neutralizing Omicron variant in the Japanese population

**DOI:** 10.1101/2022.02.23.22271433

**Authors:** Hitoshi Kawasuji, Yoshitomo Morinaga, Hideki Tani, Yumiko Saga, Makito Kaneda, Yushi Murai, Akitoshi Ueno, Yuki Miyajima, Yasutaka Fukui, Kentaro Nagaoka, Chikako Ono, Yoshiharu Matsuura, Hideki Niimi, Yoshihiro Yamamoto

## Abstract

**Introduction:** The vaccine against SARS-CoV-2 provides humoral immunity to fight COVID-19; however, the acquired immunity gradually declines. Booster vaccination restores reduced humoral immunity; however, its effect on newly emerging variants, such as the Omicron variant, is a concern. As the waves of COVID-19 cases and vaccine programs differ between countries, it is necessary to know the domestic effect of the booster.

**Methods:** Serum samples were obtained from healthcare workers (20-69 years old) in the Pfizer BNT162b2 vaccine program at the Toyama University Hospital 6 months after the second dose (6mA2D, n = 648) and 2 weeks after the third dose (2wA3D, n = 565). The anti-SARS-CoV-2 antibody level was measured, and neutralization against the wild-type and variants (Delta and Omicron) was evaluated using pseudotyped viruses. Data on booster-related events were collected using questionnaires.

**Results:** The median anti-SARS-CoV-2 antibody was >30.9-fold elevated after the booster (6mA2D, 710.0 U/mL [interquartile range (IQR): 443.0–1068.0 U/mL]; 2wA3D, 21927 U/mL [IQR: 15321.0–>25000.0 U/mL]). Median neutralizing activity using 100-fold sera against wild-type-, Delta-, and Omicron-derived variants was elevated from 84.6%, 36.2%, and 31.2% at 6mA2D to >99.9%, 99.1%, and 94.6% at 2wA3D, respectively. The anti-SARS-CoV-2 antibody levels were significantly elevated in individuals with fever ≥37.5 °C, general fatigue, and myalgia, local swelling, and local hardness.

**Conclusion:** The booster effect, especially against the Omicron variant, was observed in the Japanese population. These findings contribute to the precise understanding of the efficacy and side effects of the booster and the promotion of vaccine campaigns.

## INTRODUCTION

The effect of the vaccine against severe acute respiratory syndrome coronavirus 2 (SARS-CoV-2) gradually declines after a two-dose regimen [1-3]. For recovery of immunization, additional vaccination as the third dose boosts antibody and neutralizing responses [4]. Its effectiveness was also confirmed against the previous Variant of Concern (VOC) [5, 6]. However, the appearance of the Omicron variant, which was newly registered as a VOC in November 2022, had an impact on vaccination because it carries many amino acid changes in the spike protein [7]. Thus, the effectiveness of a vaccine booster for the Omicron variant has become a worldwide concern. The effectiveness of the booster against the Omicron variant has been reported mainly in countries and regions where the booster was previously implemented [8-12]. However, there is little evidence of the efficacy of booster vaccination against the Omicron variant in Japan because the booster was started after the appearance of the variant.

The chemiluminescence reduction neutralization test (CRNT) has been previously established to estimate the neutralization activity against SARS-CoV-2 [13-15]. Because a pseudotyped virus is used for the assay, CRNT can be handled in a biosafety level 2 laboratory and is suitable for the evaluation of a large number of samples. Using a modified method, high throughput CRNT (htCRNT), we previously reported the effect of two doses of Pfizer BNT162b2 on healthcare workers [15]. In the study, antibodies were confirmed in all participants in both the anti-SARS-CoV-2 receptor-binding domain (RBD) test (median, 2,112 U/mL; interquartile range [IQR], 1,275 to 3,390 U/mL) and the htCRNT against wild-type (median % inhibition at serum dilution, 1:100, >99.9; IQR, >99.9 to >99.9) at 2 weeks after the second dose of the BNT162b2 vaccine.

The present study aimed to follow up the humoral immunity levels after the second dose of vaccination and to evaluate the effect of the third dose of BNT162b2 vaccination, the anti-RBD antibodies and the neutralization activity against wild-type pseudotyped virus and emergent VOC, including Delta and Omicron pseudotyped viruses.

## Materials and Methods

### Specimen collection

Serum samples were collected from healthcare workers (20-69 years old) at the Toyama University Hospital. All individuals received the Pfizer BNT162b2 vaccine. A booster (the third dose) was administered after the second dose. Blood samples were collected 6 months after the second dose (6mA2D) and 2 weeks after the third dose (2wA3D). The sera were used for serological assays within 3 days of storage at 4 °C or frozen at -80 °C until further verification.

### Generation of pseudotyped viruses

Pseudotyped vesicular stomatitis viruses (VSVs) bearing SARS-CoV-2 S proteins were generated, as previously described [13]. The expression plasmid for the truncated S protein of SARS-CoV-2 and pCAG-SARS-CoV-2 S (Wuhan) was provided by Dr. Shuetsu Fukushi of the National Institute of Infectious Diseases, Japan. pCAGG-pm3-SARS2-Shu-d19-B1.617.2 (Delta-derived variant) and pCAGG-pm3-SARS2-Shu-d19-BA.1_EPE_3mut_Omi (Omicron-derived variant) were also generated, as previously described [14]. VSVs bearing envelope (G) (VSV-G) were also generated as a control. The pseudotyped VSVs were stored at −80 °C until subsequent use.

### Serological tests

The neutralizing effects of each sample against pseudotyped viruses were examined using htCRNT as previously described [14]. Briefly, serum diluted 100-fold with Dulbecco’s modified Eagle’s medium (DMEM; Nacalai Tesque, Inc., Kyoto, Japan) containing 10% heat-inactivated fetal bovine serum was incubated with pseudotyped SARS-CoV-2 for 1 h. After incubation, VeroE6/TMPRSS2 cells (JCRB1819) were treated with DMEM-containing serum and pseudotyped viruses. The infectivity of the pseudotyped viruses was determined by measuring the luciferase activity after 24 h of incubation at 37 °C. The values of samples without pseudotyped virus and those with the pseudotyped virus but without serum were defined as 0% and 100% infection (100% and 0% inhibition), respectively.

To measure the half-maximal neutralizing titer (NT_50_) values, the pooled samples made by mixing equal volumes in a single tube were serially diluted, and neutralization activity was measured in duplicates by htCRNT. The NT_50_ was defined as the maximum serum dilution that indicated >50% inhibition.

The serum concentration of the anti-RBD antibody in serum samples was measured using the Elecsys Anti-SARS-CoV-2 S immunoassay (Roche Diagnostics GmbH, Basel, Switzerland) at Toyama University Hospital. Because the upper limit of quantification was 25000.0 U/mL, measurements of >25000.0 U/mL were considered as 25000.0 U/mL for further statistical calculations.

### Vaccine-related symptoms after the third dose of vaccination

Basic clinical characteristics were arbitrarily obtained from the questionnaires, as previously described [15]. Data on the following characteristics were obtained: age, sex, local symptoms after vaccination (pain at the injection site, redness, swelling, hardness, local muscle pain, feeling of warmth, itching, and others), and systemic symptoms after vaccination (fever ≥37.5 °C, general fatigue, headache, nasal discharge, abdominal pain, nausea, diarrhea, myalgia, joint pain, swelling of the lips and face, hives, cough, and others).

### Statistical analysis

Statistical analysis was performed using the Mann–Whitney test to compare non-parametric groups. The Friedman test with Dunn’s test was used for multiple comparisons among the three paired groups. Correlations between the test findings were expressed using Pearson’s correlation coefficients. Data were analyzed using GraphPad Prism version 9.3.1 (GraphPad Software, San Diego, CA). Statistical significance between different groups was defined as P < 0.05. Data were expressed as medians with interquartile ranges (IQRs).

### Ethics approval

This study was performed in accordance with the Declaration of Helsinki and was approved by the ethical review board of the University of Toyama (approval No.: R2019167). Written informed consent was obtained from all the participants.

## RESULTS

### Antibody quantification and neutralizing activity before and after the booster

Serum samples were obtained from 648 and 565 participants, at 6mA2D and 2wA3D, respectively. At 6mA2D, the median concentration of anti-RBD antibody was 710.0 U/mL (IQR: 443.0–1068.0 U/mL) and the median htCRNT value for wild-type was 84.6% (IQR: 73.7–92.3%) (**Fig. 1 and 2A**). At 2wA3D, both the values were elevated, and the median concentration of anti-RBD antibody and the median htCRNT value were 21927 U/mL (IQR: 15321.0–>25000.0 U/mL) and >99.9% (IQR: 99.9–>99.9%), respectively (**Fig. 1 and 2A**). For VOC, median htCRNT value of Delta and Omicron was elevated from 36.2% (IQR: 19.0–52.9%) and 31.2% (IQR: 11.2–46.5%) at 6mA2D to 99.1% (IQR: 98.4–99.6%) and 94.6% (IQR: 92.4–96.3%) at 2wA3D, respectively (**Fig. 2A**).

**Figure 1.**
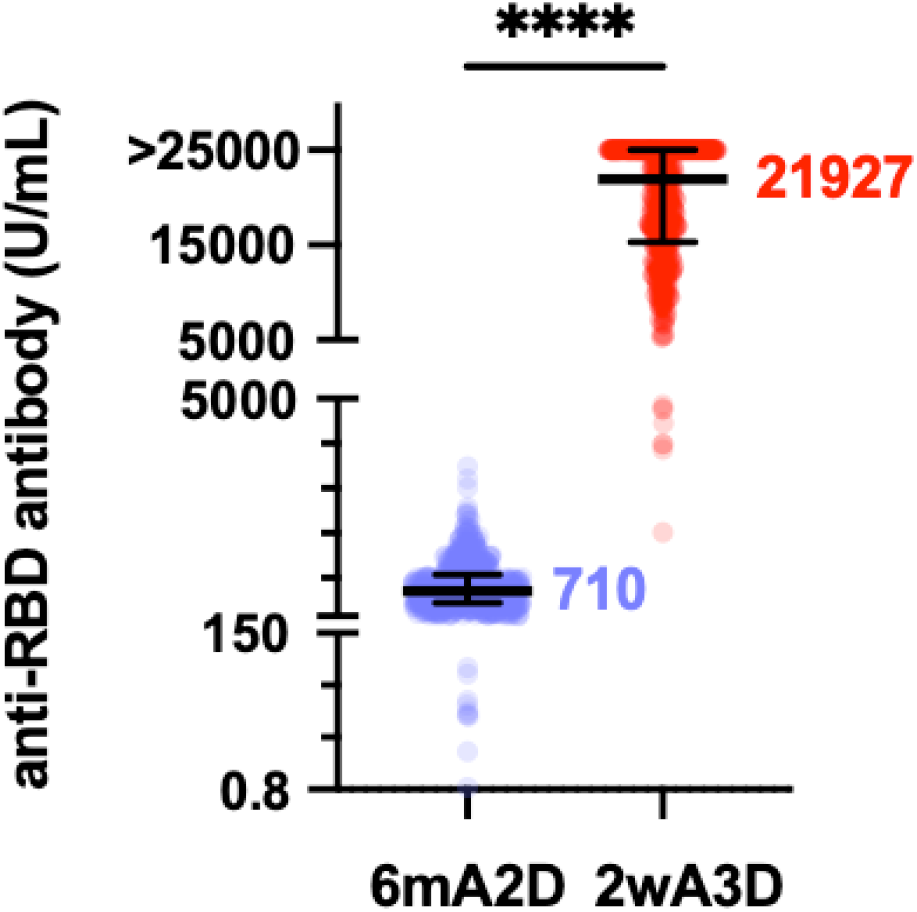
The anti-RBD antibody levels before and after the booster. Serum concentration of anti-RBD antibody at 6mA2D (n = 648, before the booster) and 2wA3D (n = 565, after the booster). Each dot represents an individual result. RBD, receptor-binding domain; 6mA2D, 6 months after the second dose; 2wA3D, 2 weeks after the third dose. ****, p < 0.0001. Bars indicate the medians with interquartile ranges.

**Figure 2.**
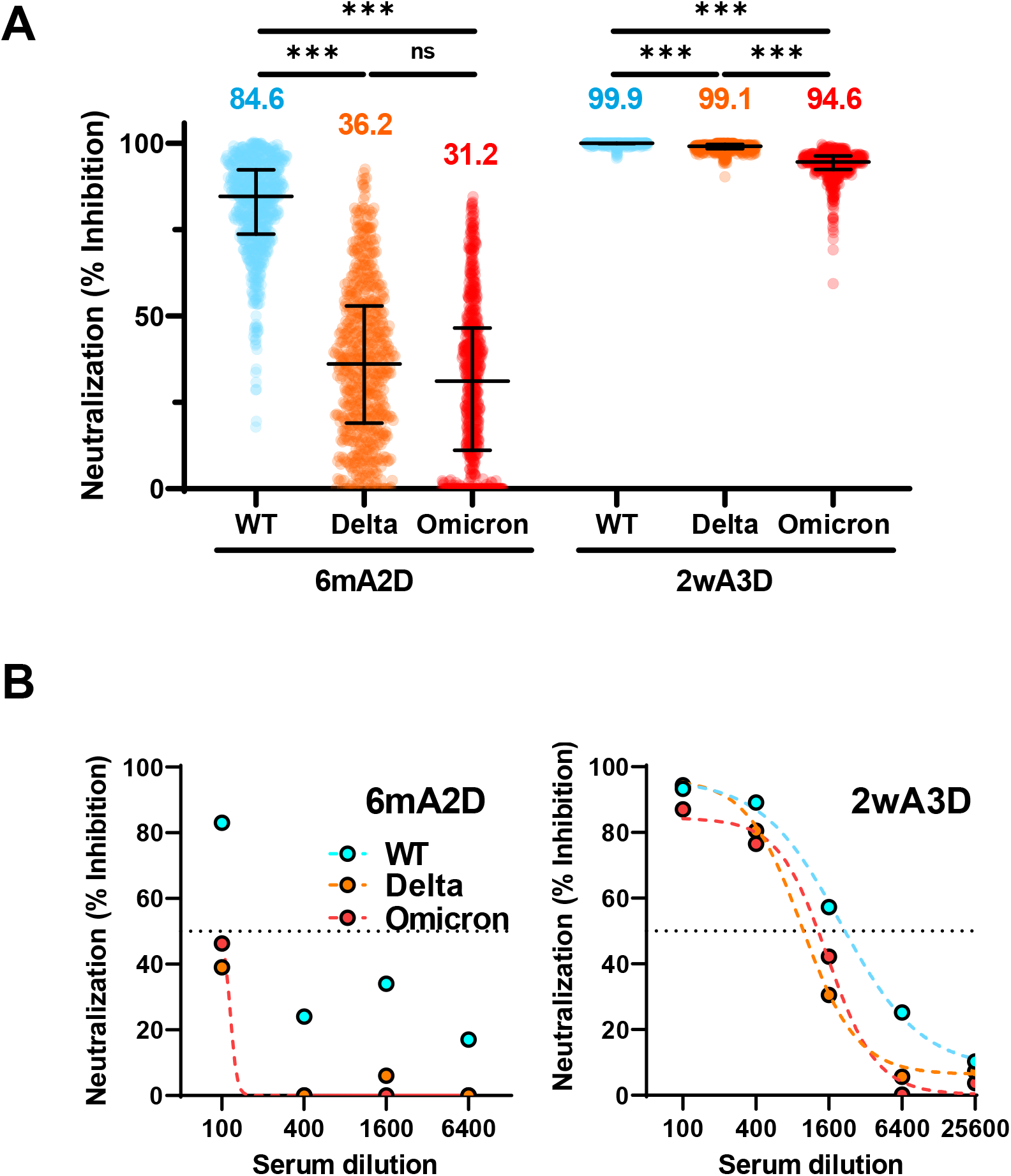
Neutralizing activity against wild-type-, Delta- and Omicron-derived variants before and after the booster. (A) Individual neutralizing activity against WT, Delta, and Omicron-derived variants at 6mA2D (n = 648) and 2wA3D (n = 565) using 100-fold diluted serum. The numbers at the top indicate the median neutralizing values of each group. (B) Neutralizing titration against WT, Delta, and Omicron-derived variants at 6mA2D (left) and 2wA3D (right) using the pooled serum. Dotted lines indicate interpolate standard curves. WT, wild-type; 6mA2D, 6 months after the second dose; 2wA3D, 2 weeks after the third dose. ***, p < 0.001. Bars indicate the medians with interquartile ranges.

The NT_50_ values of wild-type-, Delta-, and Omicron-derived pseudotyped viruses at 6mA2D were × 100, < × 100, and < × 100, respectively. NT_50_ of wild-type-derived pseudotyped virus at 2wA3D was × 1600, while that of Delta- and Omicron-derived virus was × 400 (**Fig. 2B**).

### Vaccine-related symptoms after the third dose of vaccination

A total of 510 participants provided data regarding their sex and symptoms after the third dose of vaccination (**Table 1**), and their test results were compared. There was no significant difference between males and females in anti-RBD antibody levels (female, median: 21227.0, IQR: 15249.0 to >25000.0; male, median: 22349.0, IQR: 15279.0 to >25000.0). Regarding vaccine-related symptoms after the third dose, 438 and 487 participants presented with systemic and local symptoms, respectively. The values of the anti-RBD antibody test in participants who presented with at least one systemic symptom (median: 20825.0; IQR: 15196.0 to >25000.0) were significantly higher compared with those without systemic symptoms (median: 17700.0; IQR: 11337.0 to 23630; P < 0.01), but there was no significant change in the presence or absence of any local symptoms (**Fig. 3A**). When focusing on specific systemic or local symptoms, fever ≥37.5 °C, general fatigue, myalgia, swelling, and hardness were related to higher anti-RBD antibody levels (**Fig. 3B**).

**Table 1.**
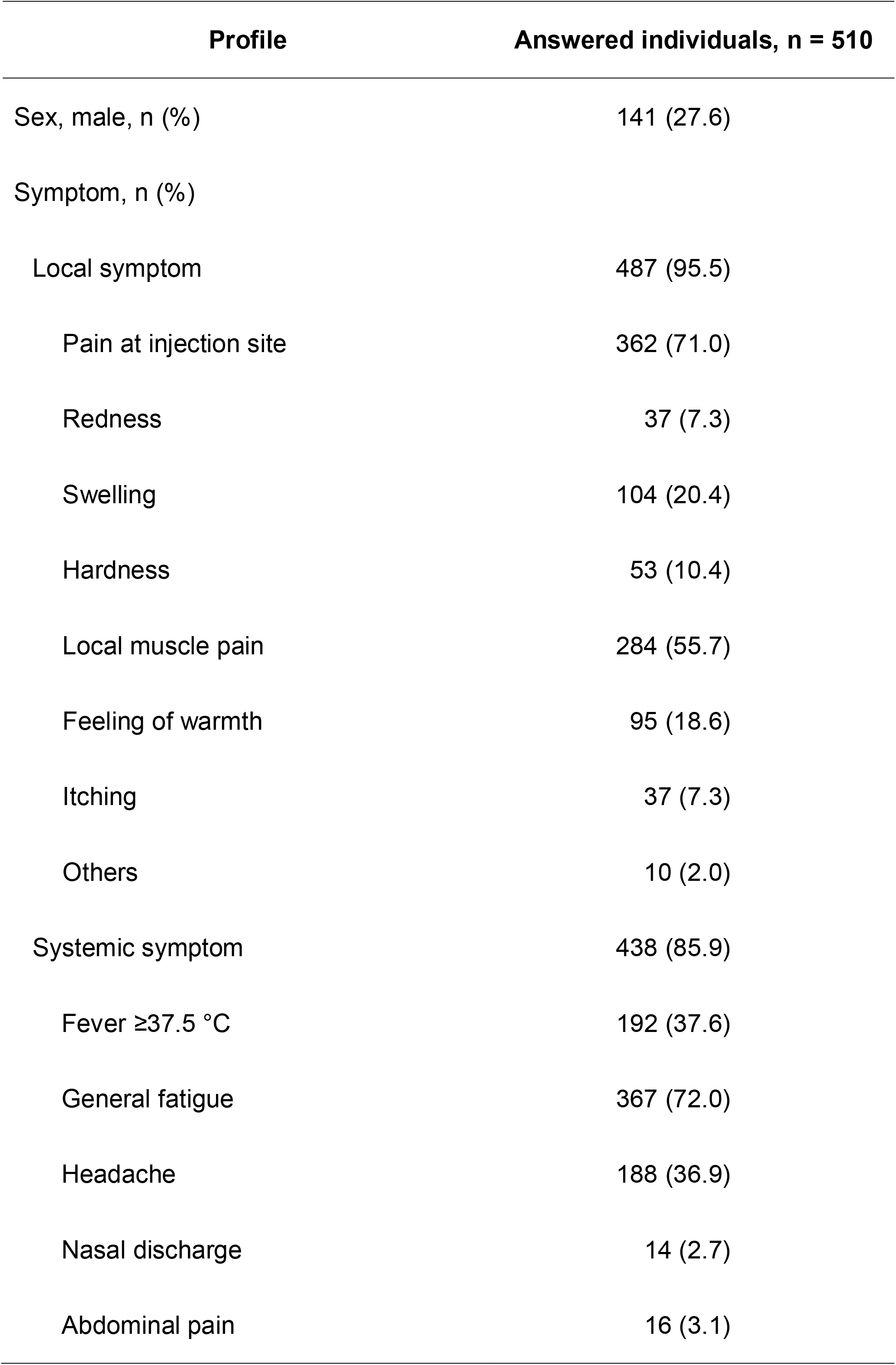
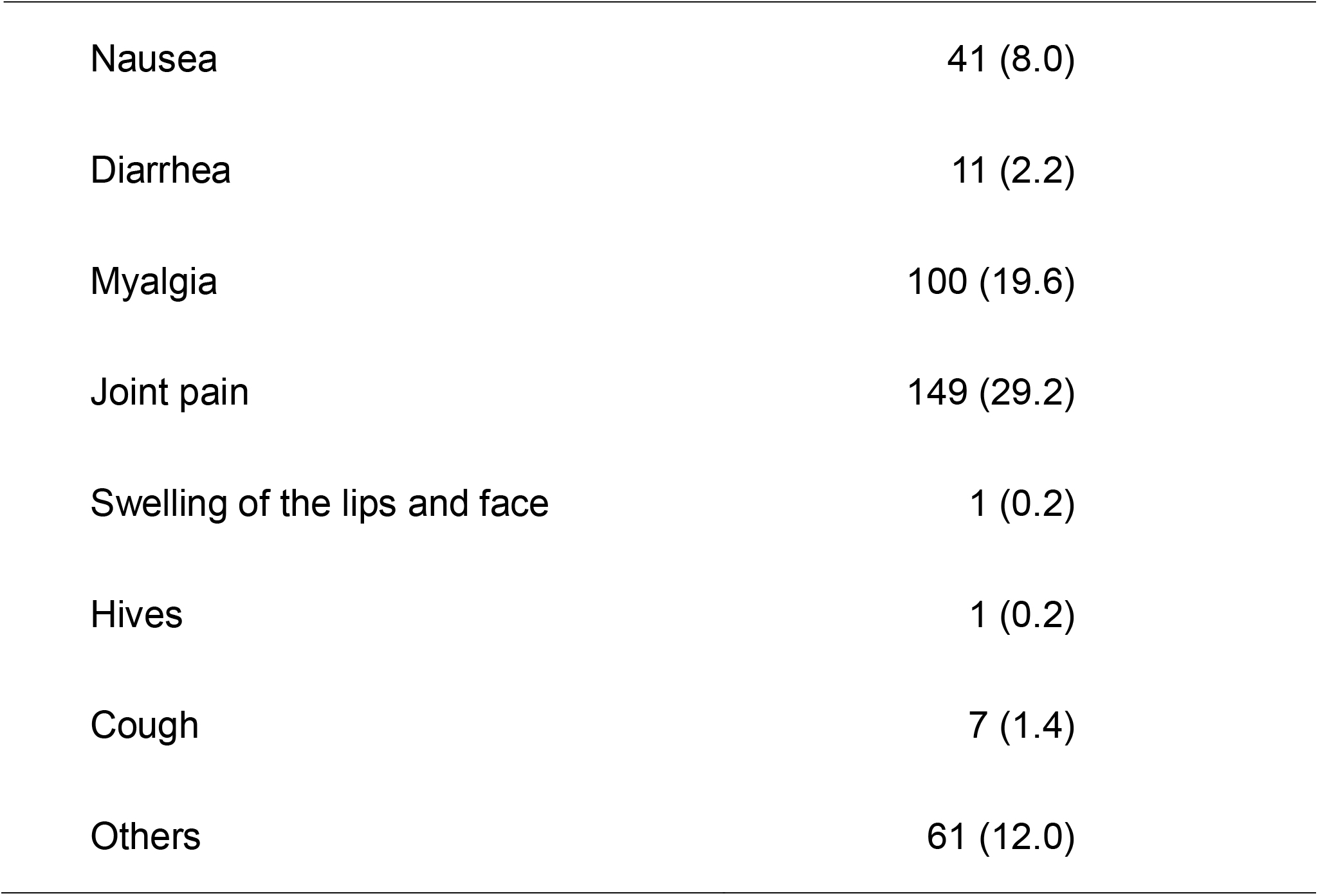
Vaccine-related symptoms after the third dose of vaccination.

| **Profile** | **Answered individuals, n = 510** |
| --- | --- |
| Sex, male, n (%) | 141 (27.6) |
| Symptom, n (%) |  |
| Local symptom | 487 (95.5) |
| Pain at injection site | 362 (71.0) |
| Redness | 37 (7.3) |
| Swelling | 104 (20.4) |
| Hardness | 53 (10.4) |
| Local muscle pain | 284 (55.7) |
| Feeling of warmth | 95 (18.6) |
| Itching | 37 (7.3) |
| Others | 10 (2.0) |
| Systemic symptom | 438 (85.9) |
| Fever $\geq 37.5$ °C | 192 (37.6) |
| General fatigue | 367 (72.0) |
| Headache | 188 (36.9) |
| Nasal discharge | 14 (2.7) |
| Abdominal pain | 16 (3.1) |
| Nausea | 41 (8.0) |
| Diarrhea | 11 (2.2) |
| Myalgia | 100 (19.6) |
| Joint pain | 149 (29.2) |
| Swelling of the lips and face | 1 (0.2) |
| Hives | 1 (0.2) |
| Cough | 7 (1.4) |
| Others | 61 (12.0) |

**Figure 3.**
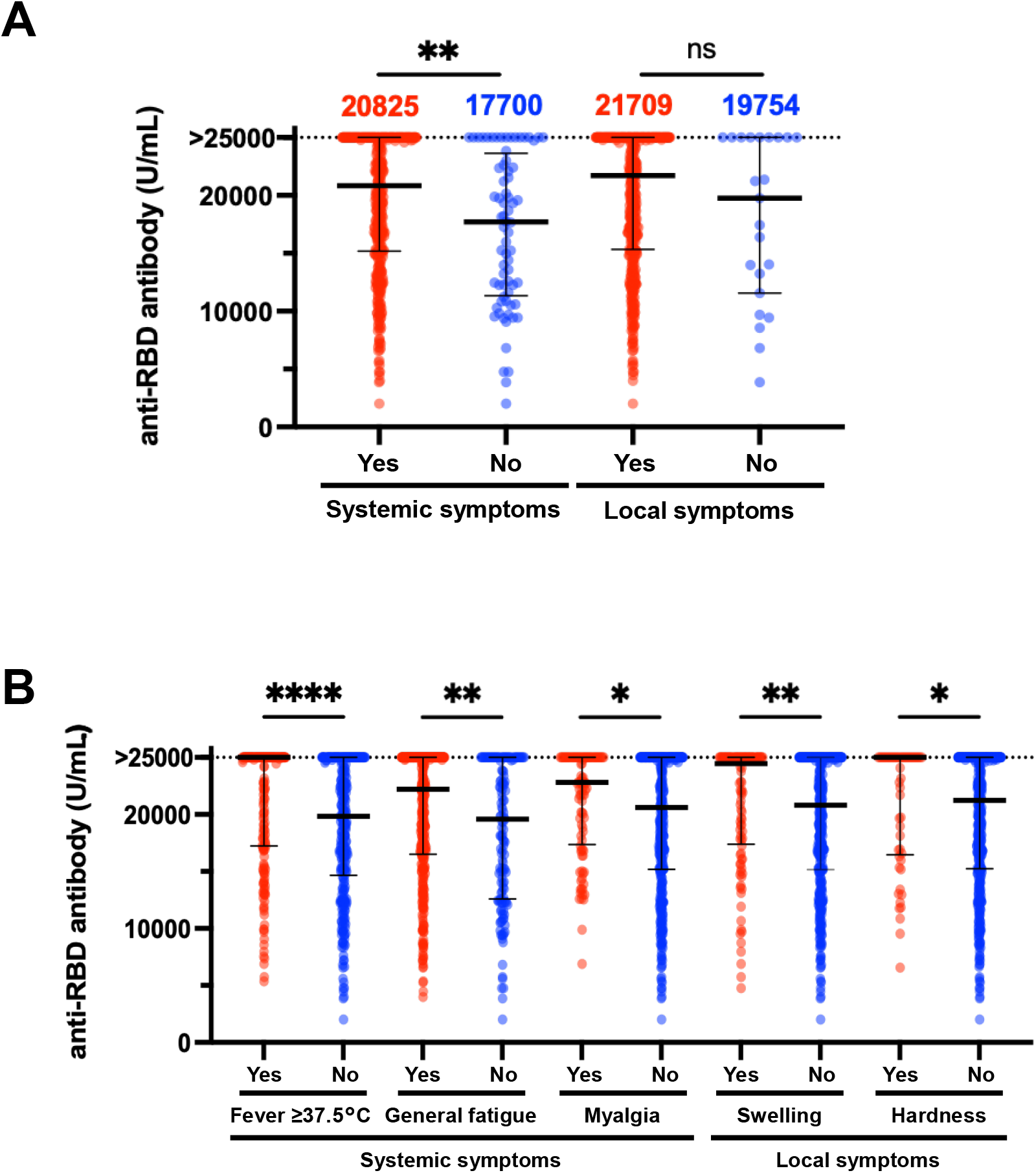
Relationship of vaccine-induced antibody levels and vaccine-related symptoms after the booster in questionnaire-answered population. (A) Anti-RBD antibody levels in individuals with systemic or local symptoms at 2wA3D (n□=□510). (B) Relationship between anti-RBD antibody levels and specific symptoms including fever ≥37.5 °C, general fatigue, myalgia, swelling, and hardness at 2wA3D (n□=□510). RBD, receptor-binding domain; 2wA3D, 2 weeks after the third dose. *, p < 0.05; **, p < 0.01; ***, p < 0.001. ****, p < 0.0001; ns, not significant. Bars indicate the medians with interquartile ranges.

## DISCUSSION

The present study showed that the booster significantly increased the amount of anti-SARS-CoV-2 antibodies and neutralizing activity in the Japanese population.

In the present study, anti-SARS-CoV-2 specific antibody levels were elevated after the booster, as previously reported [16, 17]. Compared with the anti-RBD antibody levels at 2 weeks after the two-dose regimen in our previous study (median, 2112 U/mL), the level was three-fold reduced at 6mA2D but dramatically elevated after the booster. The acquisition of antibodies is expected to prevent infection and disease progression; however, it is insufficient to determine the ability of antibodies to block viral infection and their effectiveness against variants. Because the acquisition of neutralizing antibodies correlates with coronavirus disease 2019 (COVID-19) severity [18, 19], it is important to evaluate not only the amount of antibody but also its neutralizing activity [20].

Because of a large number of amino acid changes in the spike protein, humoral immunity obtained by immunization against the Omicron variant has been a major concern compared to previous variants [7]. In the present study, the Omicron-pseudotyped virus evaded neutralization more efficiently than the wild-type- and Delta-pseudotyped viruses, as previously reported [8, 9, 11, 12, 21]. These findings are consistent with those of a study on neutralization using sera after two-dose vaccination in Japan [22].

Consistent with the results of previous reports [5, 10], our results depicted that the reduced neutralizing activity at the long interval (≥5 months) from the second dose of vaccination recovered after the booster. A previous study during a surge of COVID-19, mostly by Delta variant, indicated that the rate of breakthrough infections after the booster among healthcare workers was 0.7%, whereas that after the two-dose regimen was 21.4% [23]. For persons aged ≥60 years, the rates of confirmed COVID-19 and severe illnesses were substantially lower among those who received a booster dose of the BNT162b2 vaccine than the two-dose regimen [5]. Therefore, it is suggested that the booster, which induces stronger neutralization against the Omicron variant, can be more effective in preventing infection or severe illness than the two-dose regimen. However, little is known about the long-term effectiveness of booster therapies. A previous study demonstrated that the effectiveness against symptomatic Omicron infection peaked around 4-5 weeks after the third dose of BNT162b2 or Moderna mRNA-1273 vaccine and gradually decreased up to ≥ 12 weeks for BNT162b2 or ≥12 weeks for mRNA-1273 [24]. The neutralizing antibody titer against the Omicron variant at 6 months after the third dose of mRNA-1273 was reportedly 6.3-fold lower than that at 1 month after the booster [12]. Thus, understanding the dynamics of vaccine-mediated antibodies is still necessary.

The adverse events due to the booster were similar to those reported previously [25]. However, in Japan, limited available information limits the public’s trust and acceptance of additional dose recommendations. We demonstrated details of adverse reactions due to the booster vaccine in Japanese, and some specific local or systemic symptoms that reflect strong vaccine immunoreaction leading to higher humoral immunity. This valuable information may contribute to vaccine acceptance and relieve the fear of potential short-term side effects.

The findings in the present study are based on BNT162b2 because all participants received a planned BNT162b2 vaccination. However, similar effectiveness was also observed with other vaccines, including the mRNA-1273 vaccine [4]. The decreased neutralizing activity against Omicron after the two-dose regimen of the mRNA-1273 vaccine increased substantially after a booster dose of the mRNA-1273 vaccine [12, 26].

The limitation of this study is that the participants were limited to 20-69 years old, and the exact recovery for older and younger people who have been vaccinated is unknown. In addition, detailed interviews regarding specific underlying diseases and medications were not conducted. Thus, the elderly and those with underlying illnesses are considered to be at a higher risk of COVID-19 until their follow-up reports are available.

In conclusion, the present study showed that the booster effect, especially against the Omicron variant, was observed in the Japanese population. Since booster vaccination is now ongoing in the elderly, followed by healthcare workers in Japan, the findings contribute to the correct understanding of the efficacy and side effects of the booster and the promotion of vaccine campaigns.

## Data Availability

All data produced in the present work are contained in the manuscript

SARS-CoV-2: severe acute respiratory syndrome coronavirus 2
COVID-19: coronavirus disease 2019
VOC: Variant of Concern
CRNT: chemiluminescence reduction neutralization test
htCRNT: high throughput chemiluminescence reduction neutralization test
RBD: receptor-binding domain
6mA2D: 6 months after the second dose
2wA3D: 2 weeks after the third dose
DMEM: Dulbecco’s modified Eagle’s medium
NT_50_: half-maximal neutralizing titer
IQR: interquartile range
VSVs: vesicular stomatitis viruses

## Transparency declaration

### Data availability

The authors confirm that the data supporting the findings of this study are available within the article.

### Conflicts of interest

The authors have no conflicts of interest to declare.

### Funding

This study was supported by the Research Program on Emerging and Re-emerging Infectious Diseases from the AMED (grant number JP20he0622035) (YM, HT, and YY), the Research Funding Grant from the president of the University of Toyama (YM, NH, and YY), Morinomiyako Medical Research Foundation (HK), and Kurozumi Medical Foundation (HK).

## Acknowledgments

We thank the staff at Toyama University Hospital for their help in collecting specimens and questionnaires.

## Footnotes

1 SARS-CoV-2: severe acute respiratory syndrome coronavirus 2, COVID-19: coronavirus disease 2019, VOC: Variant of Concern, CRNT: chemiluminescence reduction neutralization test, htCRNT: high throughput chemiluminescence reduction neutralization test, RBD: receptor-binding domain, 6mA2D: 6 months after the second dose, 2wA3D: 2 weeks after the third dose, DMEM: Dulbecco’s modified Eagle’s medium, NT_50_: half-maximal neutralizing titer, IQR: interquartile range, VSVs: vesicular stomatitis viruses

## Notes

### Competing Interest Statement

The authors have declared no competing interest.

### Funding Statement

This study was funded by Emerging and Re-emerging Infectious Diseases from the AMED (grant number JP20he0622035), the Research Funding Grant from the president of the University of Toyama, Morinomiyako Medical Research Foundation, and Kurozumi Medical Foundation.

### Author Declarations

Ethics committee/IRB of University of Toyama gave ethical approval for this work.

## REFERENCES

[1] Chemaitelly H, Tang P, Hasan MR, AlMukdad S, Yassine HM, Benslimane FM, et al. Waning of BNT162b2 vaccine protection against SARS-CoV-2 infection in Qatar. N Engl J Med. 2021;385:e83.

[2] Naaber P, Tserel L, Kangro K, Sepp E, Jürjenson V, Adamson A, et al. Dynamics of antibody response to BNT162b2 vaccine after six months: A longitudinal prospective study. Lancet Reg Health Eur. 2021;10:100208.

[3] Fiolet T, Kherabi Y, MacDonald CJ, Ghosn J and Peiffer-Smadja N. Comparing COVID -19 vaccines for their characteristics, efficacy and effectiveness against SARS-CoV-2 and variants of concern: A narrative review. Clin Microbiol Infect. 2022;28:202–21.

[4] Munro APS, Janani L, Cornelius V, Aley PK, Babbage G, Baxter D, et al. Safety and immunogenicity of seven COVID-19 vaccines as a third dose (booster) following two doses of Chadox1 nCOV-19 or BNT162b2 in the uk (cov-boost): A blinded, multicentre, randomised, controlled, phase 2 trial. Lancet. 2021;398:2258–76.

[5] Bar-On YM, Goldberg Y, Mandel M, Bodenheimer O, Freedman L, Kalkstein N, et al. Protection of BNT162b2 vaccine booster against covid-19 in Israel. N Engl J Med. 2021;385:1393–400.

[6] Barda N, Dagan N, Cohen C, Hernán MA, Lipsitch M, Kohane IS, et al. Effectiveness of a third dose of the BNT162b2 mrna COVID-19 vaccine for preventing severe outcomes in israel: An observational study. Lancet. 2021;398:2093–100.

[7] World Health Organization. Tracking SARS-CoV-2 variants. 2022.

[8] Hoffmann M, Krüger N, Schulz S, Cossmann A, Rocha C, Kempf A, et al. The Omicron variant is highly resistant against antibody-mediated neutralization: Implications for control of the COVID-19 pandemic. Cell. 2022;185:447–56.e11.

[9] Dejnirattisai W, Shaw RH, Supasa P, Liu C, Stuart AS, Pollard AJ, et al. Reduced neutralisation of SARS-CoV-2 Omicron B.1.1.529 variant by post-immunisation serum. Lancet. 2022;399:234–36.

[10] Garcia-Beltran WF, St Denis KJ, Hoelzemer A, Lam EC, Nitido AD, Sheehan ML, et al. mRNA-based COVID-19 vaccine boosters induce neutralizing immunity against SARS-CoV-2 Omicron variant. Cell. 2022;185:457–66.e4.

[11] Zhao X, Li D, Ruan W, Chen Z, Zhang R, Zheng A, et al. Effects of a prolonged booster interval on neutralization of Omicron variant. N Engl J Med. 2022, doi 10.1056/NEJMc2119426.

[12] Pajon R, Doria-Rose NA, Shen X, Schmidt SD, O’Dell S, McDanal C, et al. SARS-CoV-2 Omicron variant neutralization after mRNA-1273 booster vaccination. N Engl J Med. 2022, doi 10.1056/NEJMc2119912.

[13] Tani H, Kimura M, Tan L, Yoshida Y, Ozawa T, Kishi H, et al. Evaluation of SARS-CoV-2 neutralizing antibodies using a vesicular stomatitis virus possessing SARS-CoV-2 spike protein. Virol J. 2021;18:16.

[14] Morinaga Y, Tani H, Terasaki Y, Nomura S, Kawasuji H, Shimada T, et al. Correlation of the commercial anti-SARS-CoV-2 receptor binding domain antibody test with the chemiluminescent reduction neutralizing test and possible detection of antibodies to emerging variants. Microbiolo Spectr. 2021;9:e0056021.

[15] Kawasuji H, Morinaga Y, Tani H, Saga Y, Kaneda M, Murai Y, et al. Age-dependent reduction in neutralization against alpha and beta variants of BNT162b2 SARS-CoV-2 vaccine-induced immunity. Microbiolo Spectr. 2021;9:e0056121.

[16] Hall VG, Ferreira VH, Ku T, Ierullo M, Majchrzak-Kita B, Chaparro C, et al. Randomized trial of a third dose of mRNA-1273 vaccine in transplant recipients. N Engl J Med. 2021;385:1244–46.

[17] Re D, Seitz-Polski B, Brglez V, Carles M, Graça D, Benzaken S, et al. Humoral and cellular responses after a third dose of SARS-CoV-2 BNT162b2 vaccine in patients with lymphoid malignancies. Nat Com. 2022;13:864.

[18] Lucas C, Klein J, Sundaram ME, Liu F, Wong P, Silva J, et al. Delayed production of neutralizing antibodies correlates with fatal COVID-19. Nat Med. 2021;27:1178–86.

[19] Kawasuji H, Morinaga Y, Tani H, Kimura M, Yamada H, Yoshida Y, et al. Delayed neutralizing antibody response in the acute phase correlates with severe progression of COVID-19. Sci Rep. 2021;11:16535.

[20] Garcia-Beltran WF, Lam EC, Astudillo MG, Yang D, Miller TE, Feldman J, et al. COVID-19-neutralizing antibodies predict disease severity and survival. Cell. 2021;184:476–88.e11.

[21] Muik A, Lui BG, Wallisch AK, Bacher M, Mühl J, Reinholz J, et al. Neutralization of SARS-CoV-2 Omicron by BNT162b2 mRNA vaccine-elicited human sera. Science. 2022;375:678–80.

[22] Miyamoto S, Arashiro T, Adachi Y, Moriyama S, Kinoshita H, Kanno T, et al. Vaccination-infection interval determines cross-neutralization potency to SARS-CoV-2 Omicron after breakthrough infection by other variants. medRxiv. 2022, doi 10.1101/2021.12.28.21268481:2021.12.28.21268481.

[23] Oster Y, Benenson S, Nir-Paz R, Buda I and Cohen MJ. The effect of a third BNT162b2 vaccine on breakthrough infections in healthcare workers: A cohort analysis. Clinl Microbioly and Infect. 2022, doi 10.1016/j.cmi.2022.01.019.

[24] Chemaitelly H, Ayoub HH, AlMukdad S, Tang P, Hasan MR, Yassine HM, et al. Duration of protection of BNT162b2 and mRNA-1273 covid-19 vaccines against symptomatic SARS-CoV-2 Omicron infection in Qatar. medRxiv. 2022, doi 10.1101/2022.02.07.22270568:2022.02.07.22270568.

[25] Hause AM, Baggs J, Gee J, Marquez P, Myers TR, Shimabukuro TT, et al. Safety monitoring of an additional dose of COVID-19 vaccine—united states, august 12–september 19, 2021. MMWR Morb Mortal Wkly Rep. 2021;70:1379.

[26] Doria-Rose NA, Shen X, Schmidt SD, O’Dell S, McDanal C, Feng W, et al. Booster of mRNA-1273 strengthens SARS-CoV-2 Omicron neutralization. medRxiv. 2021, doi 10.1101/2021.12.15.21267805:2021.12.15.21267805.

